# “What slows the progress of health systems strengthening at subnational level? A Political Economy Analysis of three districts in Uganda”

**DOI:** 10.1101/2023.11.09.23298302

**Authors:** Justine Namakula, Xavier Nsabagasani, Ligia Paina, Abigail Neel, Chimwemwe Msukwa, Daniela. C. Rodriguez, Freddie Ssengooba

## Abstract

There is increasing recognition that without stronger health systems, efforts to improve global health and Universal Health Coverage cannot be achieved. Over the last three decades, initiatives to strengthen health systems in low-income countries have attracted huge investments in the context of achieving the Millennium Development Goals, the Sustainable Development Goals, as well as Universal Health Coverage. Yet, health system inadequacies persist, especially at the subnational level. Our paper presents a political economy analysis featuring a three-district case study in Uganda, where district-based health systems strengthening initiatives were implemented. The study sought to understand why health systems at the subnational level are failing to improve despite marked investments.

This problem-based political economy analysis draws from a document review and key informant interviews [N=49] at the central and district levels with government actors, development partners and civil society in three purposively selected districts. Available financial data extraction and analysis were used to complement qualitative data. We found that challenges in strengthening district health systems were numerous. Themes related to financing and planning broadly interacted to curtail progress on strengthening subnational level health systems. Specific challenges included inadequate financing, mismatch of resources and targets, convoluted financial flows, as well as unwieldy bureaucratic processes. Sticky issues related to planning process-included variations in planning cycles, conflicting interests among actors, insufficient community engagement, limited decision space, and distorted accounting mechanisms.

In conclusion, the political economy analysis lens was a useful tool that enabled understanding the dynamics of decision-making and resource allocation within district health systems as well as the performance in terms of implementation of the district work plans with existing resources. Whereas it is clear that the District health teams play a big role in service program implementation, the context in which they work needs to be improved in terms of sufficient resources, setting realistic targets, widening the decision space and capacity necessary to engage with other various stakeholders and effectively harmonize the implementation of the programs. Despite playing a crucial role of compensating for local shortfalls in resources, donor resources and engagements should not happen at the cost of the subnational voice in priority setting and decision-making.

**Key messages:**

- The challenges for Health Systems Strengthening at the district level are embedded in the structural reality as well as agency interests, power-relations, and actions.
- Insufficient resources, delayed disbursement, and extreme conditional funding undercut the effectiveness of health system planning, management, performance, and accountability.
- Distorted accountability mechanisms and conflicting incentives among subnational level actors limit district health stakeholders’ decision space, displace local priorities, and contribute to community engagement strategies are not robust.
- Subnational level actors are alienated from the central and donor driven priorities and decision-making and further constrained by bureaucracies. Hence, their decision space needs amplification.
- Implementing partners should harmonize accounting and reporting mechanisms and align them to the government systems.
- Bureaucracy related to resource allocation, financial flows, and decision-making between central and district teams hinders timely implementation of services.

## Introduction

There is widespread recognition that efforts to improve global health and to achieve Universal Health Coverage (UHC) cannot be achieved without stronger health systems (1). As a result, over the last three decades, Health Systems Strengthening (HSS) in low-income countries has attracted many investments in the context of achieving the Millennium Development Goals (MDG), the Sustainable Development Goals (SDG), as well as UHC reforms (2). Yet, the goal for UHC – from “health for all by 2000” to “UHC by 2030” – has eluded us for the last 30 years, as health system inadequacy, especially at the subnational level, has persisted. In this context, both national and global actors are concerned about sub-optimal returns on HSS investments in terms of improving health system performance, resilience, and the sustainability of the capacity thereof (3).

UHC can be defined in terms of health insurance coverage and as the basket of free-of-charge services (4), but the HSS agenda underscores the role of UHC in ensuring there is benefit to all people in a defined functional zone (3), giving priority to the district and subnational efforts for health systems strengthening (5, 6). To achieve UHC through Primary Health Care (PHC)-oriented health reforms, WHO established the concept of district health systems aimed at creating a functional zone for planning and implementation of PHC services at the subnational level (7, 8), enabled with in the decentralization framework. This stimulated devolution and decentralization efforts within the health system and renewed the need for decision-makers to understand the interplay between national and subnational levels, the politics of resource allocation, and the implications of health systems reform efforts on the services that the population receives.

Despite the breadth of lessons learnt about health systems reforms and HSS, a lot remains to be uncovered. Progress has been made on analyzing health systems functions and performance through the core building blocks, health system process elements and priority outcomes (7). More recently, the literature has focused increasingly on the interactive nature of the health system blocks to generate new understandings about interactivity, dynamism, and resilience (9–12). Although acknowledged as important, less progress has been registered about the multiplicity of actors interacting to oversee and deliver services, their power and interests, and how these can be coordinated for common system goals(13, 14). Yet, these are major challenges of multi-actor capabilities, governance, and institutional arrangements that underpin health systems strengthening agendas.

In Uganda, decentralization was piloted in a few districts in 1993 as one of the hallmarks of reforms to improve service delivery across all government programs in Uganda(15). Later, in 1997, the national resistance movement passed a Local Government Act 1997(16), later amended in 2001, 2005, and 2006 (15). The Local government Act of 1997 provided a framework to rationalize the hierarchy of decentralised administrative units from village to district level (15). Local governments (LGs) hold the mandate for planning, spending and administration of programs for a defined district or municipality(16). Like in many other countries, the LGs in Uganda have healthcare and public health mandates and form the basic unit of a health system at the subnational level (17). Over the years, district health systems have become an essential locus for improving the health system to deliver PHC (18). Indeed, international organizations such as the WHO and UNICEF have underscored a renewed focus on PHC- as a vision and operational framework for achieving UHC and sustainable development goals (19). As such, district health systems have attracted investments from international development partners. Therefore, an examination of political economy factors at the subnational level, their influence on the effectiveness of investments in sub-national systems is timely.

This article is based on a three-district case study about the political economy of subnational HSS in Uganda. The study sought to understand why health systems at the subnational level are failing to improve despite marked investments. More specifically, our study set out to explore a) how do the financing and planning ecosystems function to support HSS at subnational level and the related challenges. In addition, b) what are the plausible solutions to address these HSS plans and priorities at subnational level?

## Materials and Methods

The data used in this paper are drawn from a larger study of the UNICEF-supported District Health Systems Strengthening Initiative (DHSSi) in Kenya, Malawi, Tanzania, and Uganda(20). The goal of DHSSi was to improve sub-national health management and governance as a critical component of HSS necessary to achieve UHC. DHSSi activities were focused on capacity building for evidence use in district planning, implementation, and performance management for district health management teams as well as supporting the scale-up and professionalization of health management(20). During implementation DHSSi identified several management gaps and challenges in the enabling environment that make good management practice difficult(21), and required a better understanding of the political economy at play at subnational level. In 2019, UNICEF requested a political economy analysis of DHSSi in Kenya, Malawi, and Uganda to explore these issues. The study was conducted through a collaboration between Makerere University School of Public Health and Johns Hopkins University. Findings from the sister studies in Kenya and Malawi, as well as a cross-country synthesis of the political economy analysis findings, are available elsewhere (22–24).

In Uganda, the study team implemented a multiple case study design that employed predominantly qualitative data collection methods, complemented with some secondary data extraction for financial information(21). Three districts were purposively selected in collaboration with UNICEF-Uganda office out of the ten total districts in which the DHSSi project was implemented from 2019-2022(25). The study districts were Iganga, Kiryandongo, and Isingiro(21). Table 1 summarizes the district characteristics and case selection criteria. Convenience and proximity to Kampala was also considered to facilitate data collection.

**Table 1:** Summary of district characteristics and case selection criteria.

| Characteristics | Districts |  |  |
| --- | --- | --- | --- |
|  | Kiryandongo | Iganga | Isingiro |
| Geographical location | West | East | West |
| Year of establishment | 2010 | 1975* | 1991 |
| Total population | 266,197 | 504,197 | 486,360 |
| Health systems Performance (per UNICEF baseline) % | 73.5 | 74.9 | 69.1 |
| Position on National League table 2019 out of 135 districts | 22 | 57 | 97 |

Note: \*name changed in 1980 (26-28)

### Data collection

Data collection methods included key informant interviews (KIIs), document review, and two stakeholder dissemination and validation meetings. Using unstructured thematic guide derived from the problem-driven PEA conceptual framework, the research team conducted Forty-nine KIIs between November 2020 and April 2021 with purposively selected respondents. Respondents included representatives of local and central government ministries, funding agencies, and implementing partners (see Table 2).

**Table 2:** Summary of the categories of participants and number of interviews across districts.

|  | KIIs | Participant categories |
| --- | --- | --- |
| District 1 | 14 | District Health Officers, District planners, Biostatisticians, Chief Administrative Officers, Local Council (LC5) Chairpersons, Village Health Team members, Health Unit Management committees, Health Inspectors, Health Sub-district In-charges |
| District 2 | 14 |  |
| District 3 | 13 |  |
| Central Government | 3 | Ministry of Health |
| Implementing partners | 5 |  |
| Total number of interviews | **49** |  |
| Gender |  |  |
| Female | 9 |  |
| Male | 40 |  |

The research team organised two virtual stakeholder dissemination and validation meetings. Participants included stakeholders of the health sector budget-working group, including MOH officials and other development partners, as well as selected district leaders. All interviews were conducted in English. District-level respondents participated in interviews in person. Either national level respondents participated in person or remotely, as was feasible during the data collection period, which coincided with several COVID-19 pandemic related meeting and movement restrictions. Where respondents provided their permission, the study team recorded the KIIs.

For the document review, over forty documents on HSS, guidelines for policy and planning, as well as fiscal management were pragmatically identified and reviewed. The type of documents reviewed included grey literature and reports on the implementation and evaluation of programs, government reports and policies, and some peer-reviewed articles. Financial data were obtained from the district administrations, on request.

### Data analysis

Emerging themes from the document review were integrated into the findings of the KIIs and so were findings from the data extraction. KII audio recordings were transcribed verbatim. Transcripts were reviewed by the team and analysis was conducted manually, guided by the framework used in the study design. The PEA analytical framework by Siddiqi et al 2009 (29),was used to identify the parameters that are by nature political but not necessarily explicit in the data but rather expressed through idioms, metaphors, and other expressions. Therefore, latent content analysis was a key consideration in the analytical approach. The PEA framework focuses on a specific problem or policy to better understand a challenging issue and the institutional dynamics contributing to the problem, including the broader actors and systems factors that facilitate or hinder change (30–32). Details about stages of analysis and collaborations among teams across domains have been elaborated in recently published papers emerging from this work(24).

### Ethics approval

The study received an exemption and approval from the Johns Hopkins School of Public Health (JHSPH) ethics board (IRB.no. 12934). The protocol was submitted and cleared by the Higher Degrees, Research and Ethics Committee (HDREC) of Makerere University School of Public Health (Protocol No. 890), and Uganda National Council for Science and Technology (UNCST) for approval (SS664ES).

### Ethical statement

Formal written consent was obtained from all study participants at both national and subnational level. Given the political nature of the study, participants were assured of anonymity. The study team also decided to give codes according to category of organization level that was being represented by a particular participant. For instance, terms such as “district respondent” and “IP respondent” were used. Within the text for findings, the districts were blindly given numbers 1, 2, 3, for anonymity purposes. The district name to which the number was assigned, is only known to the study team. All data was stored on a password protected computer, accessed only by the research team members.

Furthermore, Additionally, the study was conducted in the context of the COVID-19 pandemic. As a result, there were other extra standard operational procedures provided by the Ministry of Health such as social distancing, use of masks, and hand washing/sanitization, which were also strictly observed by the study team. While the study team had planned a mix of virtual and in-person interviews, respondents at the district level preferred the latter.

## Results

We structure our findings around two broad themes, which interact to curtail progress on HSS at subnational level: financing and the planning process. Under each of these themes, we highlight the main sub-themes that emerged from our political economy analysis.

### Financing

The sticky issues around financing related to insufficient amount of funds disbursed, delays in disbursement, Insufficient locally generated revenue, resource allocation priorities, mismatch between resources and targets, and duplicative bureaucracies for implementation and related perceptions among actors.

#### Insufficient resources at the sub-national level

Our analysis confirmed grossly insufficient resources allocated from the Central government to LGs, relative to the population size and expected obligations in healthcare service delivery. Several KII respondents suggested inadequate financial and non-financial resources at the subnational level as the most pervasive problem to all HSS efforts. Findings from the document review indicated that the primary reason for constantly low resource allocation from the centre was insufficient national revenues to the health sector as well as inflation (33). Indeed, a financial analysis (Fig 1) of available data on government allocations illustrated a negligible increase overtime in funding to our study districts for fiscal years 2018/19, 2019/2020 and 2020/2021.

**Figure 1:**
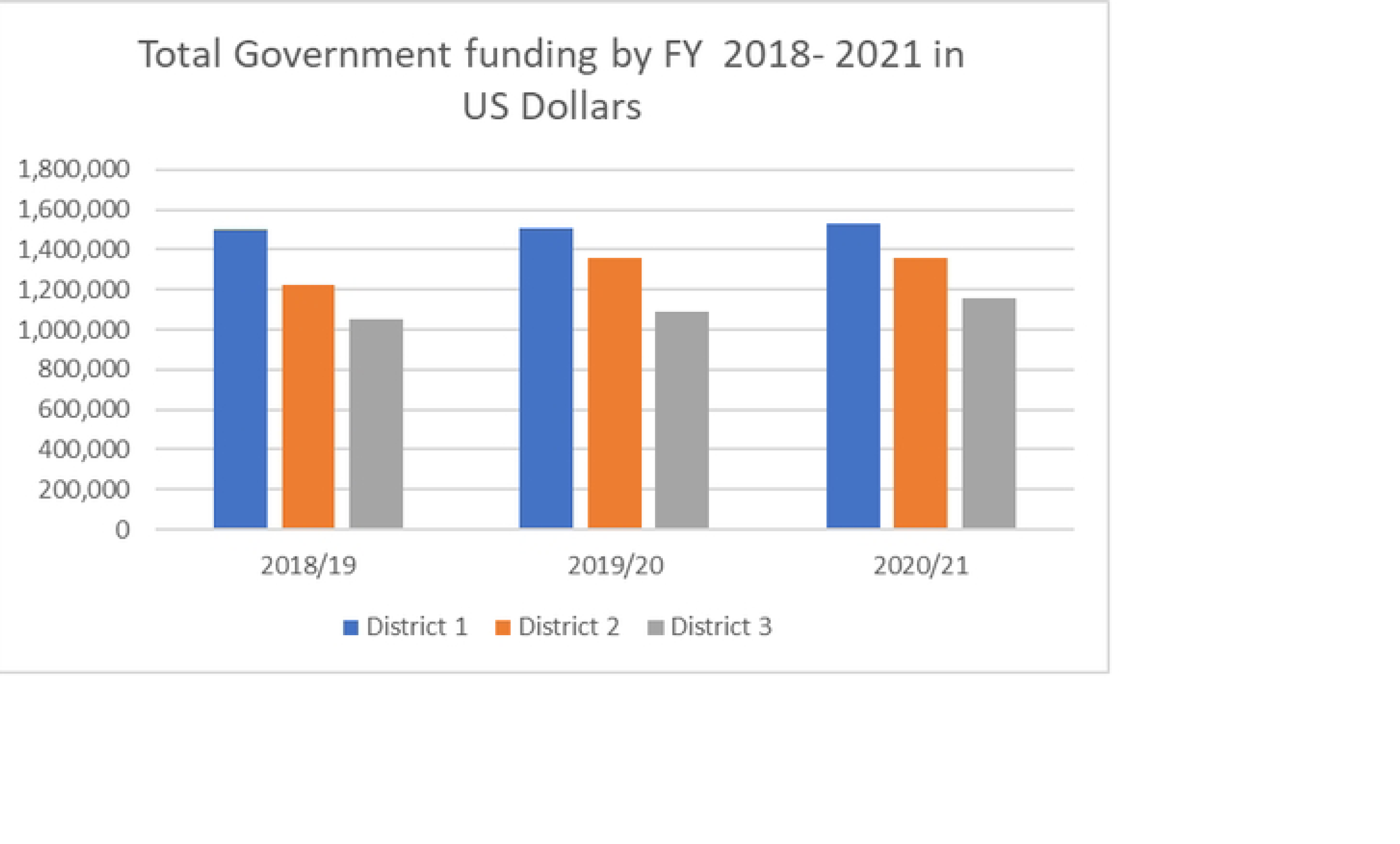
Total government funding by fiscal year (2018–2021) in US Dollars Source: Financial data extracted from study districts.

Despite the contextual differences across districts in terms of population size, location, year established, and other demographic factors, the difference in the funds allocated across the board was negligible. Moreover, the main source of funding from the centre is the Conditional Primary Health Care (PHC) allocations, which are disbursed on a quarterly basis. PHC funding covers several activities linked to centrally determined annual priorities, and is accompanied with guidelines and budget items, hence providing limited flexibility for reallocation at the district level.

Although the available data across all three districts points to stable resource allocations, most of our respondents reported a perceived reduction in financial allocations from the central government. Findings of the document review indicated the “reduction” over time was due to monetary loss of value resulting from (1) inflation, and (2) the rapid proliferation of districts and administrative units.

Further complicating resource availability, insufficient financial resources from the central government created high dependence on funding streams that have pre-determined priorities, such as donor funding whose priorities are determined at the global or national level, without consideration of specific subnational problems, priorities, and needs. According to the respondents’ reflections, the funding conditions directly affected planning and the local decision space and often at the expense of local priorities.

#### Delays in disbursement

Across all districts, respondents reported significant delays in receiving available resources, meager as they are, further delaying the implementation of district work plans. While there were reports of improvements in the timeliness of disbursements in district 2, coping strategies innovated by subnational managers from districts 1 and 2 were needed to ensure implementation of health activities. These included asking health workers to “loan their time’,” district offices taking loans/debt for the needed services and commodities from local service providers including petrol stations and stationary shops. In both cases, subnational managers deferred payments, however, in extreme situations, they mentioned that they were forced to delay or cancel the implementation of some of the activities. Respondents from district 3 never mentioned any self-innovated coping strategies, rather they leveraged on support from implementing partners.

> *“And yet getting the money is not quick […] we go ahead and do the activity on debt. […]We do the accountability and pay later. So […] requesting the people [health workers] to do the activity […] as we are processing […] we go to fuel stations and borrow the fuel then we tell the people to do the work. As for stationery, there are contracted suppliers. We issue them with Local Purchase Orders [LPOs] and they give us the stationery.” (District respondent_07)*

The financial data needed for the analysis of available budgets for specific health service delivery and technical areas was unavailable and with many missing data points. Across all three districts, there were gaps in financial data, although district 3 was worse off as it lacked data for two fiscal years. In relation to financial data from donors compared to funds from government for specific service delivery areas, there were data gaps as well. The amount of donor funding was greater than government funding for most of the technical areas where data existed (table 3).

**Table 3:**
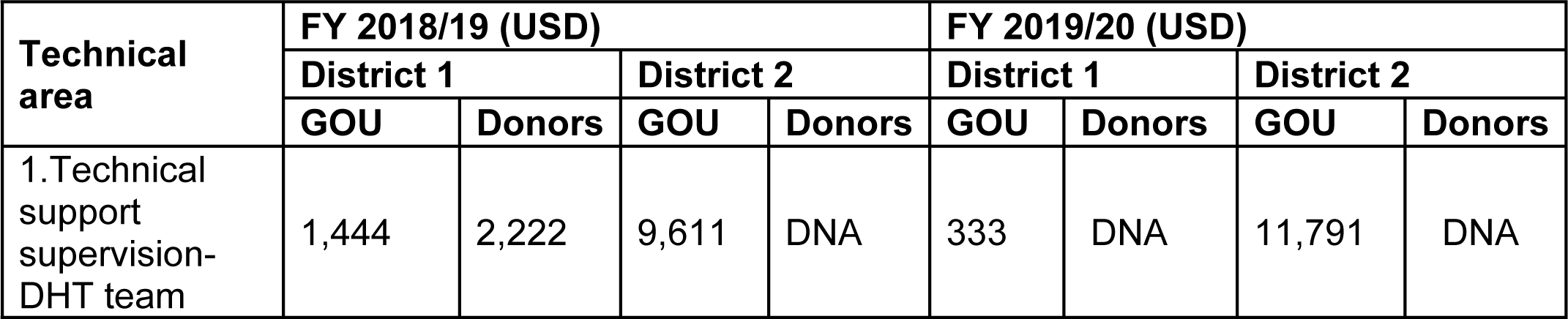
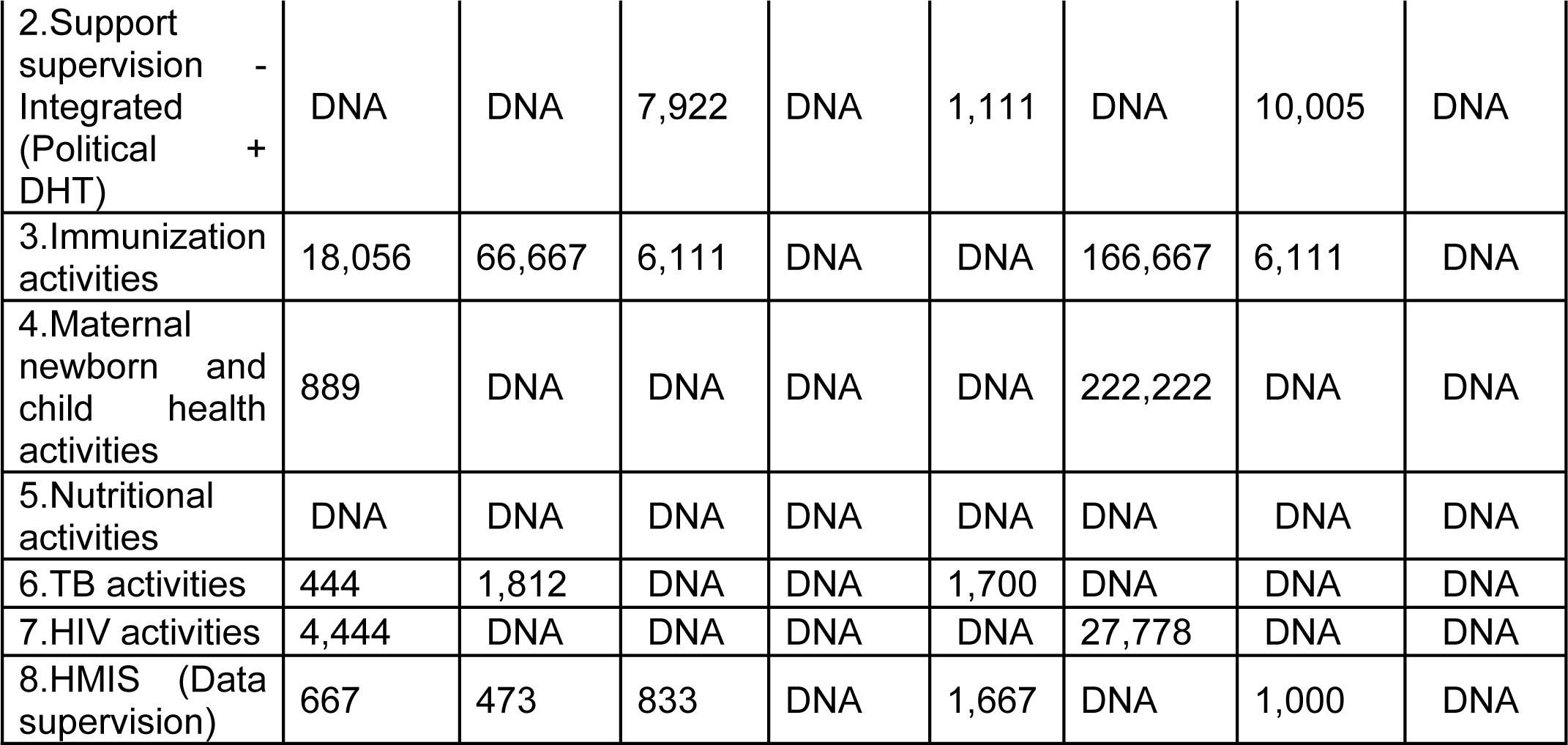
Comparative analysis of government vs donor funding by technical areas.

Note: Dollar equivalent at rate of 3600/=. DNA= Data not available, Source: financial data extraction and analysis by study team.

Further analysis of available data from district 1 and 2 showed more consistency of availability of funds for some specific programs such as immunization and maternal and child health, compared to other areas.

#### Insufficient and ‘inequitable’ Local revenue

The Local Government Act 1997 enables local governments to collect taxes as a reliable source of funding (16). The local revenue generated would ideally provide a fallback option when delays occur with central funding. Besides, the local revenue is earmarked to fund areas such as salaries for the staff not on government pay rolls. However, in addition to local tax revenues being limited, politics at the district level rendered it difficult for the health department to access such funds. For example, in one of the districts by politicians at local government level perceived the District Health Office to be “richer than other departments” because it had many externally funded implementing partners supporting activities. Therefore, monies were either siphoned by those in charge or “preserved for worse off departments,” hence hindering implementation of some operational activities for health.

> *“[…] we are not able to give a good helping hand into the health sector, given the availability of some donors in the department than any other department. So, having extraordinarily little PHC has again caused issues at the facilities because you may find a very bushy compound, facilities don’t have fencing […] (District Respondent_05)*

> *[…] As health [department] we have taken long without benefiting from it [Local revenue] so, it is as if it is not there. (District Respondent_17)*

#### Resource allocation priorities

The study found that fund holders’ priorities were biased towards greater investments in “software,” aimed at improving information technology (IT), trainings, and behavioral change communication, data for planning, report cards and community engagements for accountability. In contrast, subnational respondents reported several major “hardware” investment priorities that are needed to strengthen subnational health systems, such as infrastructure, health workforce strengthening, building maternity homes, providing more nurses and doctors, building accommodation for health workers, and increasing the stocks for medicines. Most hardware priorities were focused on infrastructural development, which, except in very few instances, tended to be ignored by both the central government and implementing partners (IPs). In a few instances, there was a mention of implementing partners focusing more on renovations and refurbishments. Nevertheless, as stated by some respondents, such hardware investment needs perpetually remain on the list of “unfunded” priorities and the system remains fragile.

> *“[…] but these [infrastructural developments] remain unfunded because there is always no money and Partners do not want to fund such things…they [IPs] reached an understanding amongst themselves that they do not want to go through capital development.” (District Respondent_ 07)*

#### Mismatch between resources and service delivery targets or expectations

The mismatch between the service delivery targets and resources also emerged as a key issue. The respondents attributed blamed this on the inadequacy of resources, coupled with overwhelming local community needs and expectations, unrealistic demands from central government, and the general dilemma of cost-efficiency (i.e., desire to do so much using minimal resources). The study found that many frustrations about HSS interventions at subnational level arise from having too grand expectations from IPs and central government despite little resources made available to LGs. As reflected by district respondents’ experiences in the case of the Measles Rubella Campaign of 2020, expectations of targets were misaligned with operational costs and realities on the ground. Unfortunately, despite the discomfort, the LGs had no choice, but to accept whatever is received and work within the limited resources. The nature of hierarchy between local government and the central government provides no room for the former to negotiate.

> *“For measles rubella, the Ministry of Health […] thought for us […] and they said that district Y, we are giving you this much and you are going to implement it this way. Of course, we wanted to tell them that it might not work. Let me give you an example. They calculated that we have 200 and something schools, but here the [District Education Officer] DEO had 938 schools [in the records]. Now, […] on the ground, we also found day care centres, which were operating without DEO’s knowledge. So, we had to work within the available budget to meet all this demand. We had to arm-twist [staff] […]” (District Respondent_04)*.

Furthermore, in relation to planning for human resources for health, one of the district managers acknowledged the mismatch of needs and targets as an apparent problem that hindered plans for recruitment and filling of staffing gaps, with district managers resorting to identifying ways of uncomfortably managing with in the minimum or leaving the status quo.

> *“[…] you cannot implement something for which you know there is no money. For example, if you have 400 health workers and they are taking 4 billion and[yet] you need 700 health workers[who] they will take 10 billion you cannot just say I want 700 health workers at least you keep on increasing, because the resource envelope is small. So […] we have not gotten funding for all the needs because the needs are far more than the resources.” (District Respondent_17)*

#### Duplicative bureaucracies for implementation and related perceptions

The study found that many donors channel resources to districts by sub-contracting IPs to implement some of the programs at sub-national level instead of utilizing existing district structures. This created a missed opportunity for supporting district capacity strengthening directly and for increasing the financial resources available locally to district leaders for implementing their work plans.

> *“[…] if UNICEF is doing work in one place, Baylor is doing work [on behalf of UNICEF]. It [UNICEF] does not work there itself. […] They do not give you cash […]” (District Respondent _13)*

At the same time, local government respondents reported frustrations and perceived increases in workload related to the separate bureaucracies and coordination introduced by implementing partner projects. Some respondents also suggested feelings of mistrust, which contributed to strained relationships between subnational level actors and IPs. For example, multiple IPs working at the sub-national level were reported to have introduced an extra layer of data, requisition and reporting structures, tools and guidelines for reporting and accountability on top of those from central government. Accordingly, respondents also noted challenges harmonizing the approaches, rules, and guidelines among diverse actors in the district, which created reporting burden on subnational governments within tight timelines. Furthermore, the general perception among district respondents was that IPs were not as transparent as expected, specifically about available budgets, which complicated planning for service delivery at subnational level.

> *“[…] they [IPs] do not want us to see their budget.” District Respondent _15)*

> *“[…] Most of them [IPs] do not disclose. They will say we are going to do this, but you may not know about the budget, how much [money] they must spend.” (District Respondent _33)*

When probed about the issue, IP respondents indicated that creating separate bureaucracies arose due to increasing concerns about local corruption, delays in accountability, and a greater emphasis on increasing value for money from donor agencies. These influenced IP practices such as not extending direct transfer of funds to districts or electronically wiring payments directly to beneficiaries or participants.

> *“We [IPs] do not give money directly to the districts […] when the activity is approved, we tell them [district team] to [go] ahead and start implementation. […] For example, for a meeting, we pay attendees via mobile [phone] money […] (District respondent_10)*

> *“But our[donor/IP] funding does not allow that [direct cash transfer to district accounts]. While we do budget support, it is in kind. We will not put money on the district account […] if it [is] the health worker who has done the activity, we send the money directly to that health worker […] (IP Respondent_19)*

#### Plausible solutions in relation to financing challenges

In response to the finance related challenges, the respondents also identified broad recommendations about what central government and donors should to address some of the challenges. These recommendations were checked and validated during the dissemination workshops and are summarised in table 4

**Table 4:** Plausible solutions in relation to financing challenges.

|  |
| --- |
| 1. The central government should expand LGs' decision space to allow them to allocate resources according to local community priorities, in line with a decentralized governance system. |
| 2. Donors and central government should empower LGs to negotiate realistic health targets, seeking to realign targets with the resources available and associated operational costs of service delivery. |
| 3. All implementing partners focusing on systems strengthening at sub-national level should prioritize "hardware" investments, e.g., infrastructure, health workforce strengthening. "Software" investments, e.g., Data for planning, IT, community engagement for accountability will gain more traction after hardware gaps have been addressed. |
| 4. Donors should directly finance health systems directly rather than engaging third-party organizations, such as IPs, which may absorb extensive financial and operational costs for HSS activities, resources that could be directly invested into district systems. In the short term, Governments and donors should consider pooling of funds to reduce waste as well as eliminating third parties' costs. |

During the validation meetings, the district respondents confirmed the findings while both the central government and IPs responded accordingly and tended to be defensive justifying what was going on instead of agreeing with the recommendations.

### The planning processes

In relation to planning and related processes, the key issues emerging from our data included conflicting interests and motivations among HSS actors at sub-national level, distorted accountability mechanisms, variations in planning cycles, shortcuts to community engagement. We expand on each of them and their interactions in the sections below.

#### Conflicting interests and motivations among actors

The study found a proliferation of actors within the subnational level planning space, with varying interests and power, crowding the decision space. These actors are located at both the district and national levels. At subnational level, actors include district level managers, technical teams, extended district management teams, administrators, political leaders, implementing partners, and the community representatives. Engagement of many actors in the local planning processes increases transactional costs and creates a congested space for decision-making. In addition, the influence of politicians at the subnational level during the planning process led to diversion of discussions and has a potential effect on outcomes for prioritization.

Despite the oversight role planned by central government agencies and the coordination role attempted by subnational managers, the influence of IPs at the subnational level cannot be underestimated. For instance, two of the study districts report receiving support for Refugees from IPs. Although the government refugee policy emphasizes integration and equal access for all, IPs were reported to exclusively prioritize refugee populations’ access to health services. This was interpreted as unfair treatment for the host communities, hence resulting in some conflict among refugees and host communities.

#### Distorted accountability mechanisms

The study identified dual track systems for accountability created by multiplicity of guidelines that undermine the subnational government decision space leading to distortions in the financing and accounting systems and a lack of mutual accountability within the health system. Even within the internal systems of the subnational governments, there were unequal relations in the distribution of resources due to power relations. For instance, the local priorities as defined by the district health management teams (DHMTs) were ignored by implementing partners in favor of priorities of the most powerful (fund holders).

Subnational level governments in general experienced challenges related to existence of many accountability rules from the central government and donors. District representatives reported penalties for returning unspent monies, as well as punishment for unmet service delivery needs relative targets set at the centre. For example, some did not receive additional funds until reporting obligations were made. This created a vicious cycle of deficient performance and inequalities among districts. Fewer innovations and rules were found to address the accountability to LGs from fund holders, however, and LGs were expected to adapt health plans in a context of frequent disbursement delays and shifting budgets.

> *“[…] actually, there is no financial year that moves in an appropriate and timely manner […] that has not happened; all the phases are delayed.” (District Respondent _12)*

> *“At the end of the fiscal year, you will be told budget cuts. So sometimes, some budget activities are not implemented. You find sometimes you started an activity, and you have already put-on funds then it comes short. That will mean you have to push it to the next fiscal year. Those have been common issues because of the budget cuts and the failure to raise adequate revenue by sub-national/government to meet at least these gaps” District Respondent_33*

Consequently, we found that the subnational level actors generally had limited decision space around implementing the district, facility, and community priorities. This was because of dependence on the centre for conditional grants coupled with pre-determined priorities with limited room for flexibility. The existence of many rules related to the planning process impeded it. For example, some respondents were concerned that the over-layering of rules created thick bureaucracies, unnecessary red tapes, and rigidity, which paralyzed decision making and ultimately affected planning and implementation at the local level.

> *“[…] Governments are a strait jacket. If you try to do creative, thinking that you are doing a noble cause, you will ‘burn your fingers’”. (District Respondent _11)*

#### Variations in planning cycles

Furthermore, respondents reported variations in planning cycles and priorities for IPs and districts, which created distortions and made harmonization with district priorities difficult. However, the district respondents noted that they continuously tolerated the situation for fear of “losing” their support to other districts.

> *“[…] the challenge we have with IPs is…their budget cycle is not the same as ours and some of them run a Calendar year, yet we run a financial year […] (District respondent _ 15)*

> *“ […] Even before we have our budget approval for the next year […] often, the district is already finalizing their budget and since we have not yet finalized our budget, we cannot commit funds to the district before the donor approves*.

> *The district cycle starts in October […] our cycle starts in July. Sometimes you find that there is a certain lag in between” (IP respondent_19)*

#### Shortcuts to community engagement

Community participation in planning and accountability is subjective to many different definitions, scope, and purposes. This ranges from community as represented by elected leaders at the district and municipalities, members of the health unit management committee, or village level residents. The rules for community participation assume that the needs of the community will shape the financial priorities at subnational level. We found that the definition and operationalisation of community participation in planning varied in study districts, many shortcuts to community participation. These included using routine health data to inform district plans, using data from previous plans, or pseudo representation of the community. As a result, the communities lost trust in the planning processes due to unmet expectations.

> *[…] you cannot start to say you can consult the whole country over certain things, you will fail; but when you look at the proxy indicators, they can help you to understand the community. (District Respondent _13)*

> *[…]Sometimes we sit and think that these health facility management committees are [very] powerful in making things work but not much. […] they cannot go beyond recommending. Moreover, their ‘recommendations’ are crafted by the medical officer or the in charge or whoever and it may not be really biting. (District Respondent _11)*

#### Plausible solutions for Planning process challenges

**Table 5:**
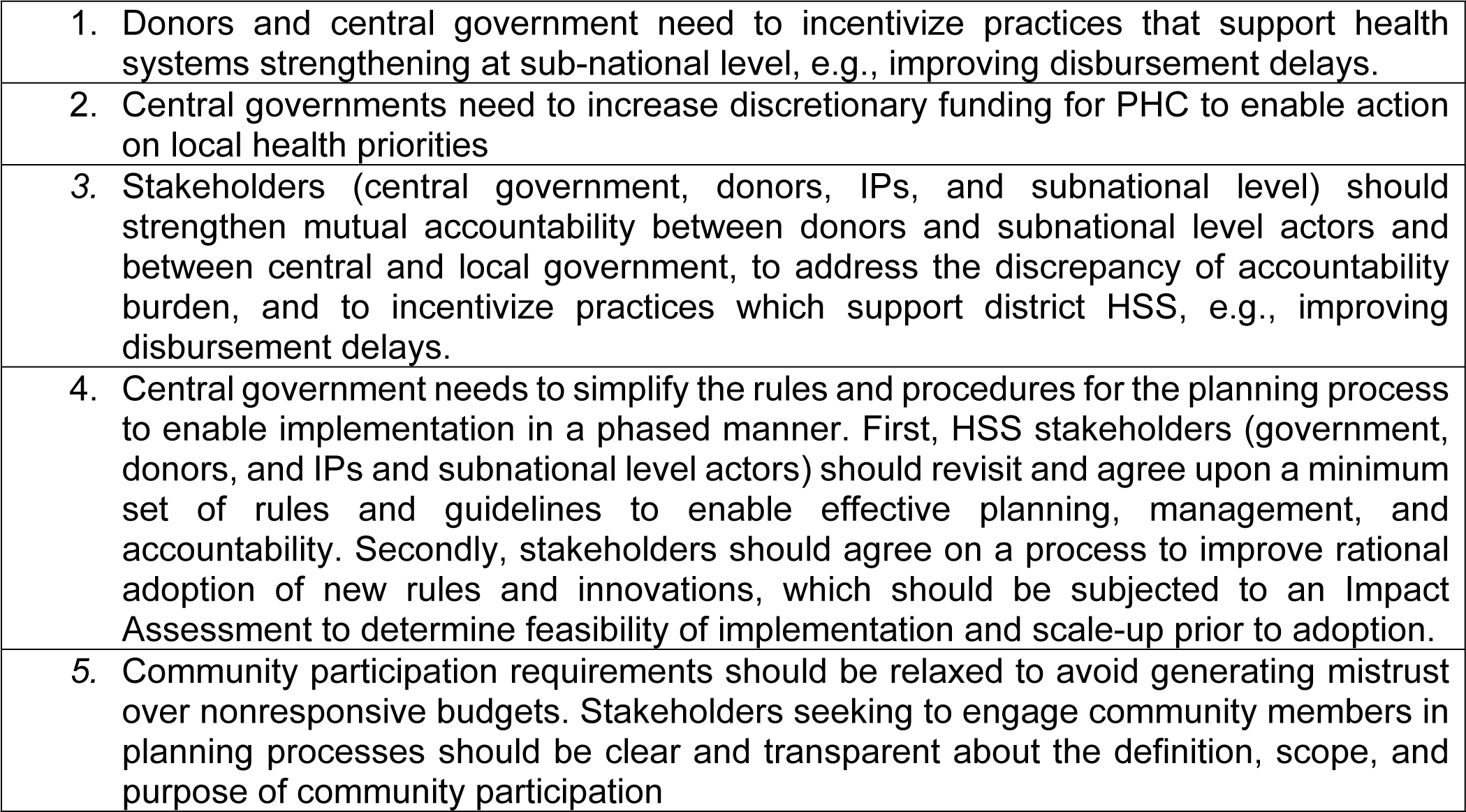
solutions for Planning process challenges.

## Discussion

The performance of subnational/local governments in the health sector has remained sub-optimal despite consensus on the importance of subnational health systems to the delivery of quality, affordable, and equitable health services, as well as despite heavy investments in HSS. The findings from our application of the PEA framework accentuate the fact that to understand a problem, you must reflect on the underlying characteristics of the problem as well as its root causes (Siddiqi et al 2009). In Uganda, we found that the problem of sub-optimal performance of the health system at sub-national level was driven by a constellation of factors that limited subnational decision space and created barriers to implementation. These include several complex dilemmas: inadequate resources as well as underspent budgets, insufficient management capacity at local level as well as duplicative bureaucracies, a proliferation of external actors engaged in Subnational government decision-making, often at the expense of engaging with local communities, and unrealized implementation plans and underutilized data, resulting in health plans and budgets inadequately responsive to community-generated priorities(21).

These health system challenges reflect, in part, an incomplete decentralization process in Uganda. While district health teams at subnational level have taken on increasing responsibility for health service delivery, fiscal and planning autonomy are yet to be achieved. Subnational governments remain dependent on the central government for funding, which often oversteps their mandate and dictates subnational government plans through conditional grants. In comparison with Malawi(23) and Kenya(22), the situation was unique to Uganda(24). The challenges further reflect the complex dynamics between a country’s bureaucracy and external funders. In Uganda, subnational level actors are similarly beholden to donors and IPs that are active in their districts. Subnational actors rely on additional funding streams from external partners to stretch health dollars. This influx of funding gives donors and IPs major influence in defining district-level priorities, however, and often deprioritizes health and creates duplicative reporting streams which place additional administrative burden on DHMTs. Streamlining bureaucratic processes and engagement to focus on shared priorities is critical but is far from being realised.

Finally, inadequate financing emerged as the main factor hindering DHMTs to advance HSS efforts and meet the ever-growing population needs and demands. The proportion of the national budgetary allocation to health sector continues to fall short of the Abuja declaration recommended target of 15% (34, 35), albeit a slight increment from 7.8% in 2019/20 (36)to 9.8% in FY 2021/22(37). This finding has implications for strategies to empower district governments with revenue as well as increased bargaining power to negotiate with funders to address local priorities.

### Study strengths and study limitations

This study is the first problem-based political economy analysis of its kind and the most recent one conducted in Uganda. Together with its sister studies in Malawi and Kenya, it richly updates the prior literature reflecting on decentralization, as well as on the consequences of this process on the health sector, in Uganda and more broadly in Eastern Africa.

Nevertheless, this analysis had several limitations. First, it was not possible to obtain all the necessary financial data to conduct a complete financial analysis as well as interpretation. Second, our study did not examine the role of the private not for profit sector, which plays a significant role filling in gaps in public service delivery in Uganda. Third, due to COVID-19 pandemic restrictions and effects prevented the study team to capture the effects on district planning fully and restricted engagement with national level stakeholders through video-interviews. Repeat interviews were not possible, so there was only limited follow-up to emerging themes. Saturation was not achieved in all themes and sub-themes, and a cross-case comparison among our case districts was not possible.

## Conclusion

In conclusion, the Political Economy Analysis lens is a useful approach to understanding why health systems strengthening interventions at subnational level in Uganda have stagnated despite massive investments. The challenges for HSS are embedded in the structural reality as well as agency interests, power-relations, and actions, which are often at the expense of the local governments’ means and decision space. An extensive examination of this reality by vote holders and central decision makers and donors will thus help to address the massive challenges highlighted in the analysis. The recommendations provided by respondents remain a wish list, because the district representatives are alienated from the centrally and donor driven priorities and decision making and further constrained by the related bureaucracies. Future research should not overlook the political economy dimensions present within subnational government units and its influence on resource allocation as well as the performance of implementing district work plans with existing resources. Development partners will continue to play a significant role in compensation for financial and technical local shortfalls. However, their support to HSS, donor resources and engagement should not happen at the cost of the subnational voice in priority setting and decision-making. The tension between national and subnational units will remain and will continue to be problematic until subnational units have a bigger say and control over the way the resources are allocated, capacity necessary to lead their programs in a truly decentralized fashion.

## Data Availability

All data relating to this study is available at Makerere University School of Public Health can be availed in conformity to institutional data governance policy. Data requests should be forwarded to.

## Acknowledgements

The COVID-19 pandemic broke out during our data collection period and increased the pressure and demands on national and local decision makers. Our appreciation goes to all the individuals who agreed to participate in this study at both national and subnational level. Special thanks to district level managers who worked tirelessly to ensure continued health service delivery for local communities. We thank UNICEF-Eastern and Southern Africa Regional Office, UNICEF country office in Uganda and Ministry of Health in Uganda for their feedback and support in disseminating findings.

We also acknowledge the commitment from the members of the study team at School of Public Health, Makerere University. Without their effort, in the face of the COVID-19 pandemic, this study would not have been possible.

